# Neurotechnology-based intensive upper-extremity supplementary training for inpatients with sub-acute stroke: A feasibility study

**DOI:** 10.1101/2023.11.18.23298626

**Authors:** Reut Binyamin-Netser, Shirley Handelzalts, Noy Goldhamer, Inbar Avni, Adi Yeshurun Tayer, Yogev Koren, Ofri Bibas Levy, Shilo Kramer, Simona Bar Haim, Lior Shmuelof

## Abstract

**Background:** Intensive and high-dose upper extremity training, concentrating on movement quality in the early phase after a stroke, can enhance motor recovery compared to standard care. Unfortunately, such programs do not exist due to limited resources, patient compliance, and administrative challenges.

**Objective:** To examine the feasibility and potential efficacy, and to evaluate the resources of an intensive technology-based upper extremity training emphasizing movement quality during inpatient stroke rehabilitation.

**Methods:** Twelve subjects with hemiparesis underwent 40 60-minute sessions over a 4-week period, in addition to standard care. The training included two game-based virtual reality platforms to practice proximal (tech 1) and distal (tech 2) movements with daily assessments.

**Results:** Eight subjects completed the entire protocol, three subjects completed 34–38 sessions, and one subject completed only 27 sessions. The mean time on each task was 35±4 (tech 1) and 37±2 (tech 2) minutes per hour. The intervention was perceived as motivating and enjoyable [Intrinsic Motivation Inventory (IMI) enjoyment and interest= 6.49±0.66 out of 7] and was not associated with pain [Visual Analogue Scale (VAS) mean= 2.00±2.32]. Subjects showed large improvements in all impairment measurements (mean FMA delta= 16.5 points and ARAT delta= 22.9 points).

**Conclusions:** The results support the feasibility of a high-dose high-intensity supplementary training protocol during sub-acute hospitalization and provide suggestive evidence of its efficacy.

## Introduction

Stroke is the second leading cause of death and the third leading cause of disability-adjusted life-years^1^. One year post stroke, patients have restricted participation^2^. It is estimated that one- to two-thirds of stroke survivors worldwide require some assistance or are fully dependent on caregivers for activities of daily living^3,4^. The most common impairment after stroke is hemiparesis^5^. Six months post stroke, only 12% of patients regain full functional recovery of the upper extremity (UE)^6,7^ These devastating effects transform the lives not only of those who experience a stroke, but also of their family and caregivers.

Effective neurorehabilitation is essential for enhancing stroke recovery^8^. Nevertheless, at three months post stroke, subjects achieve only 70% of their maximal potential recovery of the UE^9–12^. One potential way to improve recovery is to increase the dosage of treatment, which was shown to be effective for the upper extremities^13–17^. Furthermore, the timing of treatment delivery is also important, as both non-human stroke models and studies in humans demonstrate a time-sensitive period of plasticity^18^. For example, adding 20 hours of training during the acute and sub-acute phases was shown to be effective in improving UE motor recovery, whereas adding the same number of training hours during the chronic phase was not^19,20^. Nevertheless, most studies investigating the effect of increasing the dosage of treatments were conducted with chronic stroke patients (>6 months after stroke onset)^13,14^. Thus, additional studies are needed to demonstrate the effect of rehabilitating dosage on sub-acute stroke recovery.

The intensity and nature of the training program may also be important for recovery. It has been reported that patients make functional movement in only half of the therapy sessions, with 32 functional repetitions per session^21^. Furthermore, in clinical practice, rehabilitation therapies focus on the level of function (i.e., task-oriented training), rather than the level of motor impairment and quality of movement^21^. In addition, previous trials have shown limited effects, possibly due to their low doses of task-oriented therapies^22–25^. Considering these results, it was recently suggested to emphasize the quality of movement, training intensity, and enriched environment in sub-acute stroke rehabilitation^26^. One way to achieve these goals is by integrating engaging videogames in therapy. Adding these treatments to conventional therapy was shown to be effective (compared to conventional therapy alone ^16^). Thus, the integration of such high-dose and intensity treatments into the clinical environment during sub-acute hospitalization should have a beneficial effect.

However, research of UE motor intervention in the sub-acute stroke population has several challenges. A major difficulty is that stroke subjects are often withdrawn and passive, and spend extended time being sedentary^27^. Supplementary treatments should therefore be highly motivating and consider the potential low compliance of the subjects. Another challenge is related to the administrative complexity of adding substantial amounts of treatment to the schedule of the subjects, without compromising it. A third challenge is recruiting enough personnel for supplementary treatments. Lastly, reliance on videogame technologies that provide subjects with online feedback of their performance requires highly reliable technologies, but these can be easily controlled by non-technical staff. All of these aspects should be developed in the context of feasibility and implementation studies.

The aim of this study was to implement and assess the feasibility of adding 40 hours of UE training based on engaging video games that emphasize the quality of movement and intensity during the early sub-acute phase after a stroke. We specifically aimed to (1) evaluate the feasibility of adding two hours a day of an intensive rehabilitation protocol that is based on these technologies for four weeks, (2) evaluate the resources and ability to manage and implement the study and intervention, and (3) conduct a preliminary evaluation of subject responses to the intervention.

## Methods

### Subjects

Twelve subjects in the sub-acute phase of stroke (mean age: 61.67 years, SD 8.80, 3 females, mean time after stroke: 32.42 days, SD 16.45) (see Table 1 for demographic details – contact the corresponding author) participated in this feasibility study. The subjects were recruited at the Adi Negev Nahalat Eran Rehabilitation Village (Israel). The study was approved by the Regional Ethical Review Board at Sheba Medical Center, Israel (Approval Number 6218-19-SMC), and registered with ClinicalTrials.gov (NCT04737395) prior to the start of enrollment. All subjects signed a consent form before performing the experiment. Inclusion criteria were: age ≥18 years, ischemic or hemorrhagic stroke (hemispheric or brainstem) confirmed by CT or MRI, first-ever stroke or previous stroke with no UE weakness before the second incident, 1 week ≤ time after stroke onset ≤ 6 weeks (mean=32.42 days, SD=16.45), active shoulder flexion of at least 20° and partial wrist and/or finger active movement (due to a limitation of the technologies used in the intervention), Fugl–Meyer Assessment (upper extremity) score < 58), and the ability to provide informed consent. Subjects were excluded if they had: a painful shoulder limiting an active forward reach, severe spasticity or non-neural loss of range of motion, or cognitive or communication impairments as determined by the clinical team and unstable medical conditions.

All subjects were included in the final analyses. All enrolled subjects were after a first stroke (no recurrent strokes in the sample).

All data produced in the present study are available upon reasonable request to the authors.

### Intervention

The aim of the program was to increase the amount of practice of the UE using customized game-based platforms that included immersive, challenging, and rewarding virtual environments. In each session, subjects had to achieve an explicit goal, while receiving online feedback and rewards for goal attainment. The intervention was composed of 120 minutes of UE training a day (divided into two therapy sessions of 60 minutes), and was given for 4 weeks, for a total of 40 hours. Each session was staffed with a 1:1 PT/OT-to-patient ratio.

We used two platforms: one for the training of proximal arm (shoulder and elbow) movements (tech 1) and another for training of distal arm (wrist and fingers) movements (tech 2).

#### MindPod™ Dolphin (tech 1) for proximal arm training

The subjects used a custom-designed immersive animation-based experience named MindPod Dolphin (which is based on patented technology licensed from KATA at Johns Hopkins University to MSquare Healthcare Inc., a MindMaze Group Company, Switzerland; (Figure 1A – contact the corresponding author)^16^. In this game, the subject controls the swimming of a virtual dolphin by moving his/her shoulder and elbow joints and performing complex exploratory movements. In the game, the dolphin swims through different ocean scenes with multiple task goals such as chasing and eating fish, escaping a shark, and jumping out of the water. This game was designed to promote the movement quality of the shoulder and elbow in all planes (3D movements) (shoulder abduction, adduction, flexion and extension, and elbow flexion and extension) throughout the active ranges of motion. The game utilizes a real-time motion capture system. The playing paretic arm of the subjects was supported by a passive mechanical exoskeleton vest (ekso UE, Ekso Bionics, San Rafael, USA) (Figure 1B - contact the corresponding author). The degree of support was provided by the exoskeleton using springs with different tension levels. The amount of spring tension needed was adjusted for each subject to allow active movement of the shoulder to at least 90° of flexion. The support of the vest allowed the subjects to play with their paretic arm in an extended active range in all directions. The arm weight support level was titrated as the subject progressed through the sessions by reducing the level of support or removing the vest. No active assistance was given by the clinician during the intervention. The game is designed in a way that every level is a bit more difficult than the one before. Each level has a goal (e.g., to eat a specific number of fish), and when the goal is obtained, the game is over and the following game level begins.

#### Hand Tutor (tech 2) for distal arm training

The HandTutor system (MediTouch, Tnuvot, Israel; Figure 1C - contact the corresponding author) consists of an ergonomic wearable glove and dedicated software with games that allow practice of active wrist movements, grip control, and finger individuation in a virtual environment. Subjects progress through games, while adjusting the range of motion that is being practiced, according to their abilities. Subjects perform flexion and extension movements of their fingers or wrist separately or simultaneously. In each session, the clinician (OT or PT) can choose which movements to work on, while emphasizing grip abilities and finger individuation. In one game, for example, subjects control a fishing rod up and down by flexing and extending an individual finger to catch swimming fish. The finger range of movement is set before the game starts. If the task is easy for the subject, the clinician increases the difficulty level of the game by making the fish move faster or by adding another control dimension, such as adding the movement of the wrist on top of the fingers in a manner that the fingers control the fishing rod movement (up or down) and the wrist controls the boat movement (left or right).

Training on both training platforms was conducted while the subjects were sitting on a plastic bath chair with back support and no hand support. The legs of the chair had an anti-slip rubber covering. In both platforms, a physical or occupational therapist was present throughout each session and provided verbal and tactile feedback to ensure high-quality movements (without compensation and with the instructed body part only) and exploration of the full workspace (in tech 1). Each session was dedicated to a single platform. At least three sessions per week were practiced on each platform.

Subjects continued with their regular rehabilitative routine that included daily physical and occupational therapy sessions and speech therapy if needed, as well as group work, hydrotherapy, and gym workouts (at least 3 treatments per day) during weekdays (Sunday to Friday). Subjects were also encouraged to work on cardiovascular fitness during the program, as well as to use their paretic upper extremity in daily activities. Outcome measures were collected throughout the intervention period on a session-by-session basis. To evaluate the potential effectiveness of the intervention, we collected data/measures at baseline (pre training/intervention), immediately post training/intervention and at 12 (±14 days) and 24 (±14 days) weeks post stroke.

### Outcome measures of feasibility

Adherence rates to study procedures were documented by the therapists for each session. Time on task (ToT, in minutes) was measured in each session using a stopwatch. The distance that the arm reached on the task (meters) was measured by tech 1. Game levels and the amount of weight support were documented by the therapist in each session, and of rehabilitation sessions outside of the intervention (usual care) during the study period.

Pain level was monitored using the Visual Analogue Scale (VAS) at the end of each session, and before and after the entire intervention. The VAS is a self-report measure consisting of a 10-centimeter line with a statement at each end representing one extreme of pain intensity (no pain and pain as bad as it could possibly be). The subject gave an indication with a pen mark on the line corresponding to their present pain level ^28^.

Exercise intensity was measured using the Rating of Perceived Exertion (RPE), where subjects subjectively rated their level of exertion during exercise (1=did not put an effort at all to 10=put in an extreme effort)^29^.

Participation was measured using the Pittsburgh Rehabilitation Participation Scale (PRPS), a clinician-rated instrument to assess the patient’s participation in therapy using effort and motivation estimates by the therapist)^30^. This measure was taken at the end of each practice session.

Motivation was measured using the IMI, a multidimensional measurement intended to assess subjects’ subjective experience related to a target activity in laboratory experiments. The instrument assesses subjects’ interest/enjoyment, perceived competence, effort, value/usefulness, felt pressure and tension, and perceived choice while performing a given activity. Motivation was assessed through rating the agreement with proposed statements using a Likert scale (1=not at all true, 7=very true)^31^ the end of the intervention period, the IMI was assessed, and adverse events and problems/difficulties related to intervention equipment (software, hardware, the vest, etc.) were documented by the therapists throughout the study period.

4.2% of data were missing from the RPE report, 5.00% from the PRPS report and 9% from the VAS report. The missing data were due to the failure of clinicians to fill in the forms.

### Outcome measures of potential efficacy

A Fugl–Meyer Upper Extremity (FM-UE) assessment was used to assess motor impairment^32^. The FM-UE has been proven to show good reliability, validity, and sensitivity to post-stroke motor changes^33^

The Action Research Arm Test (ARAT) was used to assess motor activity using 19 tests of arm motor function, both distally and proximally (grasp, grip, pinch, and gross movement)^34^. The ARAT demonstrates high reliability, and has been found useful in prior studies evaluating stroke subjects across a wide spectrum of impairments. The test shows good validity, as well as sensitivity to spontaneous and therapy-related gains after a stroke^35^

The Stroke Impact Scale (SIS) hand domain, version 2.0 was used to assess disability and health-related quality of life using a questionnaire^36,37^.

### Statistical analyses

All analyses and statistical calculations were performed using SPSS 21 (IBM SPSS statistics). Paired sample t-tests were performed between the outcome measures before and after the intervention.

The data that support the findings of this study is available on request from the corresponding author, [RBN].

## Results

The intervention was conducted on subjects with stroke from a wide range of ages (from 44 to 71), education (from 0 years of learning to 15 years), and medical conditions (obesity, schizophrenic disorder, hyperlipidemia, hypothyroidism, essential hypertension, depression) (see Table 1).

### Training adherence and time on task

The mean number of delivered sessions was 38 (SD, 4). Eight out of our 12 subjects finished 40 sessions. Subjects 3 and 11 did not finish due to COVID-19, Subject 4 had poor motivation at the end of the intervention, and Subject 12 had medical issues that prevented him from finishing the sessions (see Table 2). All subjects had pre- and immediate post-intervention assessments (100%). A total of nine patients completed the 12-week post-intervention assessment (75%), and six completed the 24-week post-intervention assessment (58.3%). Three subjects did not yet reach the time of assessment, and the others failed to attend. The mean time on task (ToT) was 35 minutes per one-hour session [SD 2.44, 33 (SD 4.35) for tech 1 (Figure 2A) and 37 (SD 2.09) for tech 2) (Figure 2B)]. For tech 1, total arm distance was also measured, reaching a mean distance of 246 meters (SD 87.35) (Figure 3). In addition to the intervention, all subjects received an average of 2.7 standard care rehabilitation hours a day. In total, they received an average of 54 hours of standard care rehabilitation during the intervention period.

**Figure 2:**
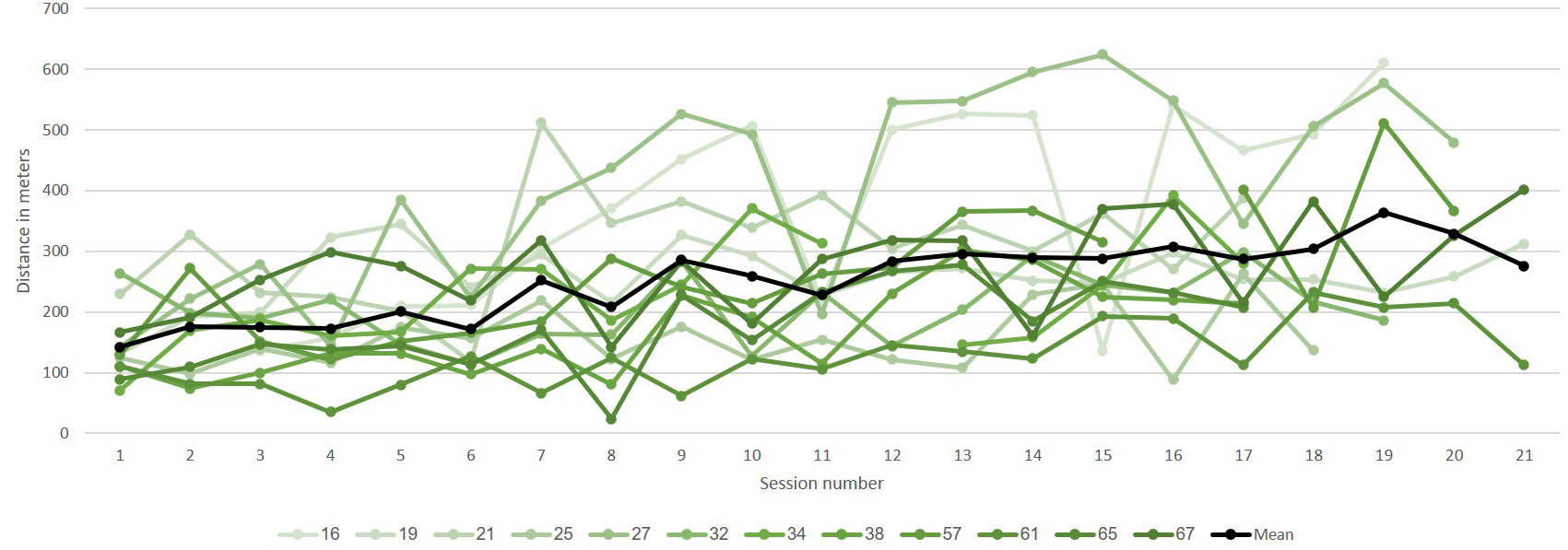
Total time on task. **A.** Tech 1 (shoulder and elbow movement practice) **B.** Tech 2 (wrist and fingers movement practice). The data is represented for each of the sessions. All 12 subjects are represented. Each subject is represented by a different line. The average of all subjects represented by a black line.

**Figure 3:**
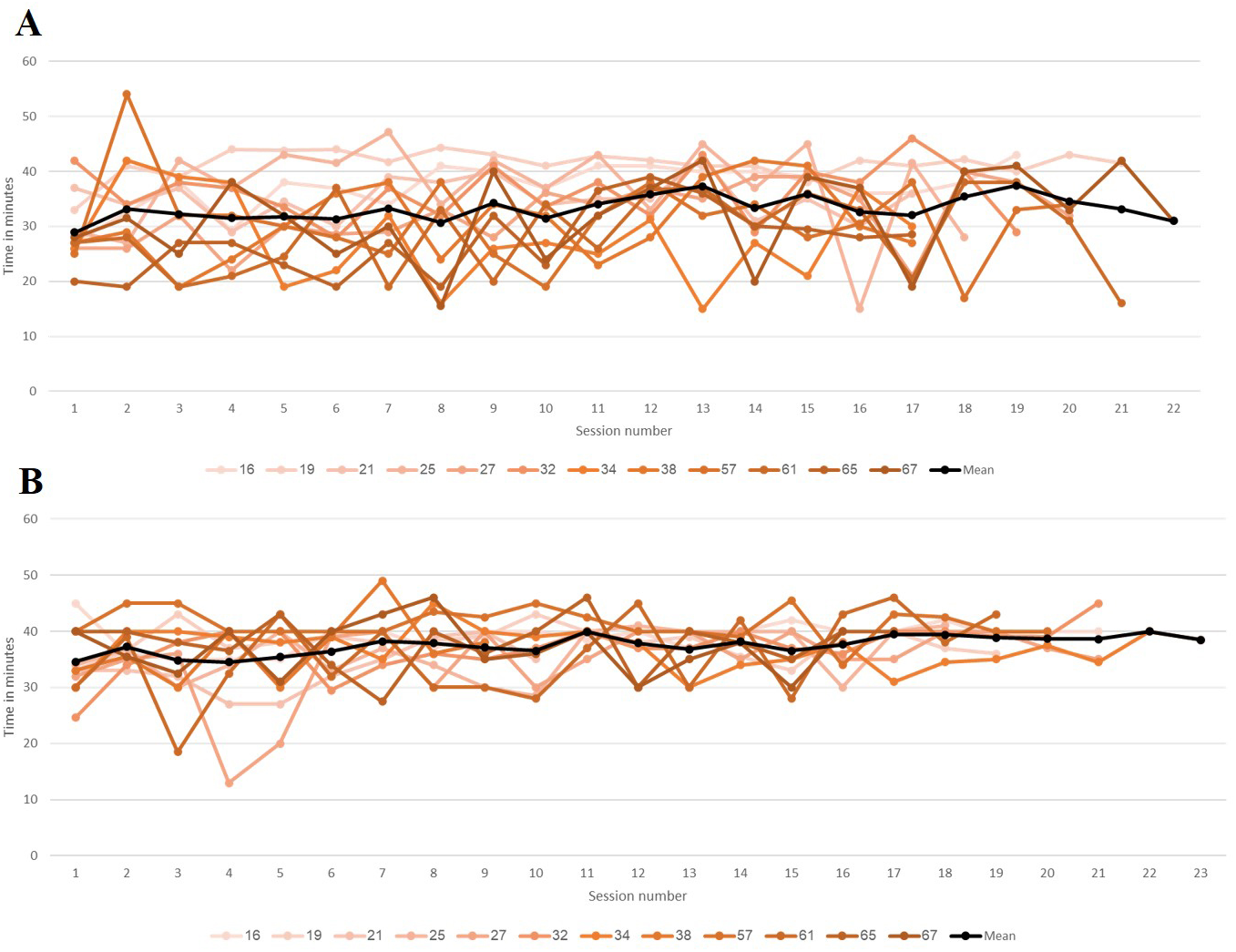
Total arm distance for tech 1 (shoulder and elbow movement practice) for each of the sessions. All 12 subjects are represented. Each subject is represented by a different line. The average of all subjects represented by a black line.

**Table 2:** Training attendance and Time on Task (ToT)

| Subjects | Total intervention sessions | Mean TOT Tech 1 (Minutes) | Mean TOT Tech 2 (Minutes) |
| --- | --- | --- | --- |
| 1 | 40 | 37 (4) | 39 (2) |
| 2 | 40 | 41 (2) | 38 (3) |
| 3 | 27 | 35 (4) | 33 (4) |
| 4 | 36 | 37 (8) | 36 (4) |
| 5 | 41 | 32 (6) | 34 (7) |
| 6 | 40 | 36 (5) | 37 (4) |
| 7 | 40 | 28 (8) | 37 (3) |
| 8 | 40 | 31 (7) | 37 (6) |
| 9 | 40 | 31 (8) | 41 (3) |
| 10 | 40 | 29 (7) | 35 (7) |
| 11 | 34 | 27 (6) | 37 (4) |
| 12 | 38 | 31 (8) | 37 (5) |
| **Mean (SD)** | **38 (4)** | **33 (4)** | **37 (2)** |

The effort level that was reported by the subjects was high (RPE, see Methods). The mean effort subjects reported was 7.14 (SD 2.18) (out of 10) [7.38 (SD 2.21) for tech 1 and 6.90 for tech 2 (SD 2.21)]. This suggests that subjects were motivated to put a lot of effort into each session. Participation assessment by the clinicians was also high (PRPS, see Methods). The mean participation of subjects was 5.42 (SD 0.49) [5.45 (SD 0.49) for tech 1 and 5.40 (SD 0.51) for tech 2]. Pain was measured using the VAS (see Methods). This measure was taken after each session to determine if the training was associated with any pain. The mean VAS subjects reported was 2.00 [SD 2.32, 2.39 (SD 2.73) for tech 1 and 1.61 (SD 1.86) for tech 2]. This suggests that pain was generally low.

Motivation was assessed at the end of the intervention (IMI, see Methods). The mean score in each category was: Interest/Enjoyment – 6.49 (SD 0.66), Perceived competence – 6.18 (SD 0.80), Perceived choice – 5.42 (SD 1.39), and Pressure/tension – 1.75 (SD 0.97). This suggests that subjects enjoyed the intervention, felt that they could do it, and that joining the intervention was their choice. They did not feel pressured during the intervention sessions.

No adverse events were recorded during the intervention sessions.

### Feasibility of operation

The intervention hours were scheduled before and after the clinical day: at 7–8 am and 3–4 pm to minimize the effect of the intervention on standard care (three treatments per day), and to secure clinicians for the intervention. This decision led to extension of the treatment day for the enrolled subjects, and it was dependent on good cooperation between the ward management staff who had to first prepare each patient in the morning and make sure they had eaten breakfast before starting the treatment day. Another challenge raised by this decision was that even though the interventions were conducted on a single subject at a time, a group of 6–7 clinicians was needed to completely fill the intervention schedule.

### Motor outcomes

The scores of the outcome measures before and after intervention are shown for each subject in Table 3. The average FMA recovery was 16.50 points. This difference is significant (t_(11)_=-5.58, p<0.001) and higher than the mean clinical important difference (MCID) (9–10 points)^38^. The average ARAT recovery was 22.92 points. This difference is also significant (t_(11)_=-6.05, p<0.001) and higher than the MCID (12–17 points)^39^. The average SIS Function and SIS Recovery change were 1.23 points and 23.33 percent, respectively – another significant difference (t_(11)_=-5.01, p<0.001; t_(11)_=-3.77, p=0.003, respectively). The outcome measures that were obtained at the end of the intervention were largely stable at 12 and 24 weeks after stroke: the average FMA was 49.25 and 49.33 points, respectively (in comparison to the average of 49.75 points after the intervention). The average ARAT scores were 39.87 and 34.67 points, respectively (in comparison to the average of 39.0 points after the intervention). The average SIS Function was 3.43 and 2.87 points, respectively (in comparison to the average of 2.98 points after the intervention). The average SIS Recovery was 65.83 and 71.67 percent, respectively (in comparison to the average of 71.25 percent after the intervention). Generally, subjects maintained their level of motor function 12- and 24-weeks post stroke.

**Table 3:** Stroke subjects’ measurement scores.

|  | FMA Motor<br>score Pre | FMA Motor<br>Change | ARAT score<br>Pre | ARAT score<br>Change | SIS<br>Function Pre | SIS<br>Function<br>Change | SIS<br>Recovery<br>Pre | SIS<br>Recovery<br>Change |
| --- | --- | --- | --- | --- | --- | --- | --- | --- |
| 1 | 41 | 21 | 24 | 32 | 2.6 | 1 | 30 | 40 |
| 2 | 27 | 9 | 2 | 10 | 1 | 0.8 | 20 | 60 |
| 3 | 25 | 3 | 5 | 0 | 2.4 | 0.2 | 85 | -5 |
| 4 | 18 | 26 | 3 | 29 | 1 | 0.2 | 45 | 25 |
| 5 | 25 | 28 | 0 | 49 | 1.2 | 1.2 | 10 | 40 |
| 6 | 40 | 12 | 36 | 18 | 2 | 2.2 | 50 | 10 |
| 7 | 33 | 10 | 26 | 14 | 1.4 | 1 | 30 | 20 |
| 8 | 38 | 8 | 12 | 19 | 2.6 | 1.8 | 80 | 20 |
| 9 | 36 | 27 | 30 | 27 | 1.4 | 1.4 | 15 | 55 |
| 10 | 30 | 32 | 11 | 36 | 1.2 | 3.2 | 80 | 0 |
| 11 | 54 | 3 | 36 | 14 | 2.4 | 0.6 | 60 | 15 |
| 12 | 32 | 19 | 8 | 27 | 1.8 | 1.2 | 70 | 0 |
| **Mean** | **33.3** | **16.5** | **16.1** | **22.9** | **1.7** | **1.2** | **47.9** | **23.3** |
| **SD** | **9.5** | **10.2** | **13.5** | **13.1** | **0.6** | **0.8** | **27.0** | **21.5** |

## Discussion

This study demonstrates the feasibility of an intensive supplementary sub-acute training protocol of 40 sessions. In the majority of cases, the protocol was implemented successfully. Incomplete implementation was due to reasons that were unrelated to the intervention protocol. The proportion of ToT from the total time was above 60%. Participants found the intervention to be motivating, enjoyable, and engaging.

### Intensive upper extremity training in sub-acute stroke patients is feasible

We showed that it is possible to add two hours of upper extremity intensive treatment per day during hospitalization. This feasibility has two components: it shows that subjects in the sub-acute stage can endure two additional hours of intensive training a day, and that the medical system can deliver this training. The following elements are recommended for program success: (1) Administrative support – this program requires the full cooperation of the rehabilitation ward staff to prepare the patient on time (prioritizing them), and to organize their treatment schedule in a way that considers factors such as meals, rest periods, and the mandatory number of usual care treatments per day. (2) Engagement of clinicians and patients – a sufficient number of clinicians should be trained to administer the intervention treatments to ensure a smooth flow of treatments (without cancelations). Coordination of the clinical staff is also important for information exchange regarding the participating patient. (3) Reliable and attractive technologies – the technologies we chose to use in this intervention were reliable, highly motivating, enjoyable, and challenging. We conclude that while we showed here that subjects can endure intensive rehabilitation, evaluation of the resources and ability to manage and implement the study and intervention is essential.

### Feasibility outcome measures

We expected to reach 2/3 of the total time dedicated for each training session in ToT (40 min per 60 minutes), similarly to the expected ratio in a standard therapy session. However, we reached only 35–37 minutes per hour for a number of reasons such as preparation time, technology maintenance, and patient fatigue. Integrating the intervention within the standard care schedule may increase the proportion of ToT.

Notably, compared to other studies that investigated intervention protocols in the sub-acute phase after a stroke, we managed to give the most intensive protocol^19,20,22–25,40^.

In addition to the potential benefits of early intervention^19,20^, the ability to provide an intensive intervention protocol in the sub-acute phase during hospitalization is highly cost effective since it does not require the maintenance of external programs such as those that are used in the chronic phase after a stroke^13,14^.

### The motor benefits of the intervention

Substantial improvement was reported in all motor outcome measures. This improvement was larger in comparison to other reported recovery measures^13,14,41^. A new study showed that adding 30 hours of upper extremity training resulted in better motor improvement than standard of care alone^16^. This study showed an improvement of 12.5–13.4 points in the FMA (in comparison to 16.5 points of improvement in our study) and 13.4–14.7 points in the ARAT (in comparison to 22.9 points of improvement in our study). We, therefore, recommend studying the efficacy of a supplementary intervention protocol of at least 40 hours.

### Implementation recommendations

Implementation of the intensive treatment could be improved in different manners: the efficacy and feasibility of the intervention could be enhanced by increasing the number of patients per clinician (OT or PT), so that multiple subjects can practice at the same time. Practice can also be administered by an assistant of an OT or PT (not requiring a senior therapist). Moreover, it may be possible to develop an implementation model where treatment is given as part of the rehabilitation day, and not during off hours.

Another suggested avenue is to expand this protocol in time and to include a telerehabilitation component. Although this type of rehabilitation is relatively new, it has been shown to be feasible^41^. Therefore, we suggest further studies regarding the implementation of high dosage and intensity with a such a component. Ultimately, the efficacy of this intervention compared to standards of care and to increased standards of care dosage should be studied in randomized controlled trials.

## Conclusion

Intensive upper extremity training is practical and well received by subjects with sub-acute stroke. Successful implementation depends on the clinical setting, technologies, and resources. We call for studying the implementation of this protocol on multiple subjects at a time and during clinical days, and for randomized control trials to study the efficacy of the intervention. Our preliminary results suggest that sub-acute intensive rehabilitation may improve motor recovery after a stroke.

## Data Availability

All data produced in the present study are available upon reasonable request to the authors

## Acknowledgments

We thank Dr. Oren Barzel for his clinical assistance. We thank the reviewers for their constructive comments. This work was supported by the Israeli Science Foundation grant 1244/22 to LS.

